# BCR::ABL1 evaluation in bone marrow is no more predictive of TFR success than from peripheral blood

**DOI:** 10.1101/2025.09.01.25334821

**Authors:** S.D. Patterson, A. Gottschalk, C. Hayden, R. Young, A. Hair, H.G. Jørgensen, H.W. Wheadon, J.F. Apperley, I. Glauche, R.E. Clark, M. Copland, I. Roeder

**Affiliations:** Paul O’Gorman Leukaemia Research Centre, School of Cancer Sciences, College of Medical, Veterinary and Life Sciences, University of Glasgow, Glasgow, UK; Institute for Medical Informatic and Biometry, Faculty of Medicine, Technische Universität Dresden, Dresden, Germany; SIHMDS, North West London Pathology, London, UK; Robertson Centre for Biostatistics, School of Health and Wellbeing, College of Medical, Veterinary and Life Sciences, University of Glasgow, Glasgow, UK; Centre for Haematology, Imperial College London, London, UK; Department of Molecular and Clinical Cancer Medicine, University of Liverpool, Liverpool, UK

**Author notes:** These authors contributed equally (S.D. Patterson, A. Gottschalk, M. Copland, I. Roeder). **Competing Interests Statement** M.C. has received research funding from Cyclacel and Incyte, is/has been an advisory board member for Novartis, Incyte, Ascentage, Jazz Pharmaceuticals, Pfizer and Servier, and has received honoraria from Astellas, Novartis, Incyte, Pfizer, Janssen and Jazz Pharmaceuticals. J.F.A has received research funding from Incyte and Pfizer, is/has been an advisory board member for Ascentage, Incyte, Novartis and Terns, and has received honoraria from Incyte and Novartis. The remaining authors have no disclosures.

## Abstract

**Background:** Despite the excellent survival for chronic myeloid leukemia (CML) patients treated with tyrosine kinase inhibitors (TKI), most CML patients report poorer quality of life due to TKI side-effects. In the DESTINY trial (NCT01804985), CML patients halved their TKI dose for 12 months prior to TKI cessation, with 24 months of follow-up off therapy. Enrolled patients (n=174) were in stable deep molecular remission (DMR; BCR::ABL1^IS^ <0.01%) or major molecular response (MMR; BCR::ABL1^IS^ <0.1%), assessed in peripheral blood (PB) following more than 3 years of TKI treatment. Molecular recurrence-free survival (MRFS) at 3-years was 72% (DMR) and 36% (MMR), indicating PB BCR::ABL1^IS^ burden at TKI de-escalation/cessation is predictive for recurrence. As shown previously, BCR::ABL1 kinetics during dose de-escalation are predictive for molecular recurrence after TKI stop. However, it has not been determined if BCR::ABL1 RT-qPCR in bone marrow (BM) improves risk prediction compared to PB.

**Objectives:** We compared the predictive value of BCR::ABL1^IS^ measurements at the time of dose de-escalation between PB and BM with respect to molecular recurrence, including assessing if the inclusion of a BM measurement at time of dose de-escalation improved the predictive power of BCR::ABL1 kinetics during de-escalation.

**Methods:** With written informed consent, PB and BM samples were collected and BCR::ABL1^IS^ was measured using RT-qPCR. Of 107 patients evaluated, 42 had molecular recurrence and 65 maintained TFR. To describe BCR::ABL1 kinetics during dose de-escalation, we applied linear regression. Estimated individual intercept and slope parameters were used within a logistic regression model to check for their predictive value for molecular recurrence.

**Results:** The median log10[BCR::ABL1^IS^] at time of dose de-escalation was higher in patients who experienced recurrence in both BM and PB (BM: −2.69 for recurrence vs. −4 for TFR; PB: −2.78 for recurrence vs. −2.98 for TFR). Based on samples with detectable log10[BCR::ABL1^IS^], transcript levels were moderately correlated between PB and BM (Spearman’s r=0.6). Using logistic regression, log10[BCR::ABL1^IS^] values from either BM or PB taken at time of de-escalation were similarly predictive of TFR success at 36 months post de-escalation (BM: OR=1.8; PB: OR=2.6). Moreover, using intercept and slope as characteristics of the treatment kinetics during dose de-escalation jointly as independent, PB-based predictors (OR_intercept_=13.8, OR_slope_=1.5) led to a similar conclusion and allowed for classification of patients into three risk groups: low, high, unclear. We identified 54 patients (50%) with low risk, 20 patients (19%) with high risk, and 33 patients (31%) with unclear risk. Considering TFR success, patients in the low-risk group show 87% MRFS 24 months after TKI stop, whereas all high-risk patients had molecular recurrence within 16 months of trial entry.

**Conclusion:** No difference in the predictive power of a single measurement taken prior to dose de-escalation using either PB or BM samples could be shown. However, predictive power was enhanced by determination of slope and intercept of sequential PB BCR::ABL1 samples obtained during TKI de-escalation. Overall, BM evaluation prior to treatment cessation was not more predictive than PB of TFR success, however using BCR::ABL1 kinetics in PB during TKI de-escalation enabled classification of patients for risk of molecular recurrence.

## Introduction

Continuous tyrosine kinase inhibitor (TKI) treatment provides rapid disease control in almost all patients with chronic phase (CP) chronic myeloid leukemia (CML), leading to near-normal survival [1, 2]. However, most patients report poorer quality-of-life due to TKI-related side-effects [3-6]. Some of the CP-CML patients achieving deep molecular responses (DMR; BCR::ABL1^IS^ ≤0.01%) can stop their TKI, often after many years of therapy, and successfully maintain major molecular response (MMR; BCR::ABL1^IS^ ≤0.1%), termed “treatment-free remission” (TFR) [7]. Longer durations of TKI treatment and DMR are associated with improved rates of successful TFR; however, a reliable prediction of individual TFR success is not possible.

The DESTINY trial [8] enrolled CP-CML patients in stable DMR or MMR, following ≥3 years of TKI treatment (see Suppl. Information). Each patient’s TKI dose was reduced by 50% of the respective standard dose for 12 months (de-escalation phase) followed by TKI cessation (stop phase). Patient outcomes were determined 24 months post-TKI cessation as either ‘TFR’ or ‘recurrence’, defined as BCR::ABL1^IS^ > 0.1% at the first of two sequential measurements > 0.1%. At 24 months post-TKI cessation, the rates of molecular recurrence-free survival (MRFS) were 72% and 36% in the DMR and MMR groups, respectively. This de-escalation approach reduced TKI-related side effects and costs whilst maintaining efficacy, and further demonstrated that BCR::ABL1^IS^ kinetics in peripheral blood (PB) during TKI de-escalation are predictive for molecular recurrence after TKI stop [9].

Here, we aimed to determine if the evaluation of BCR::ABL1 in bone marrow (BM) at the point of TKI de-escalation is more predictive of TFR than PB evaluation and whether existing prediction models could be improved by combining PB and BM measurements.

## Materials and Methods

Mononuclear cells (MNCs) were isolated from BM and PB immediately prior to TKI de-escalation (timepoint ‘0’). Of the patients enrolled in DESTINY (n=174), we conducted paired analyses of PB and BM on a subset using available samples (n=107), in which BCR::ABL1^IS^ was determined by qRT-PCR (see Suppl. Fig. 1, Suppl. Tables 1&2, Suppl. Information).

During TKI de-escalation, the molecular response was monitored monthly in PB. Linear regression modelling was applied to describe changes in PB BCR::ABL^IS^ during TKI de-escalation [9]. Using a minimum of three sequential detectable BCR::ABL1^IS^ values, the regression parameter, i.e., slope and intercept, were individually determined for all eligible DESTINY patients. In addition, BCR::ABL1^IS^ values from sequential PB samples during TKI de-escalation were compared with single BCR::ABL1^IS^ measurements taken prior to TKI de-escalation from either BM or PB (Suppl. Figure 2).

## Results

Patient demographics are shown in Suppl. Table 2. The median BCR::ABL1^IS^ value at trial entry was significantly higher in both BM and PB in patients who experienced recurrence (Figure 1A). However, BCR::ABL1^IS^ was not detectable in the BM, but detectable in PB for 42 patients (39.3%), while it was only undetectable in both BM and PB for 6 patients (5.6%) or undetectable in PB and detectable in BM for 6 patients (5.6%), suggesting PB sampling may be more sensitive (Figure 1B). Using only samples with detectable BCR::ABL1^IS^ values (n=53; 49.5%), transcript levels were moderately correlated between PB and BM (Figure 1B). Furthermore, we observed a statistically significant trend, with lower median BCR::ABL1^IS^ levels at trial entry associated with longer times to recurrence, in both BM and PB MNCs (Suppl. Figure 3). This indicates that higher BM BCR::ABL1^IS^ levels prior to dose de-escalation are associated with a shorter time to relapse, in parallel with previous findings based on PB BCR::ABL1^IS^ alone.

**Figure 1.**
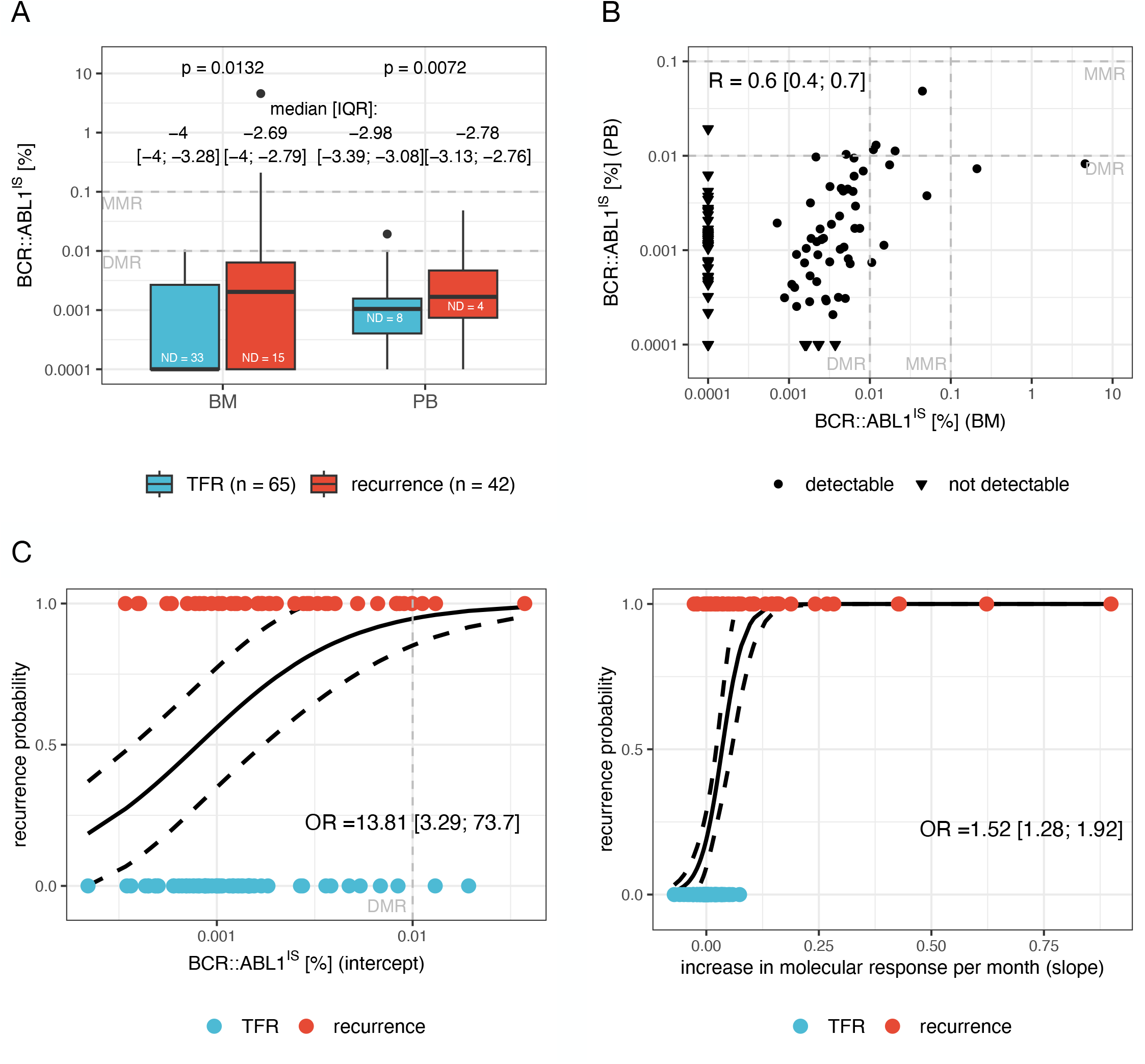
BCR::ABL1^IS^ measurement from peripheral blood (PB) is more predictive of TFR outcome than from bone marrow (BM). **A:** Box plots showing the median (black line) and interquartile range [IQR] (box) for molecular response levels (c.f. Suppl. Information) detected in BM (left) and PB (right) MNCs immediately prior to TKI dose de-escalation, stratified by outcome (maintained TFR: blue; molecular recurrence: red) 24 months post TKI cessation. Samples for which the molecular response was below the limit of detection are denoted as ‘not detectable’ (ND). *p*-values are derived from Mann–Whitney U-tests, comparing the median molecular response between TFR and recurrence patients for BM and PB measurements, respectively. **B:** Molecular response values as detected in BM and PB MNCs immediately prior to dose de-escalation. For correlation analysis, ND values were excluded and the Spearman’s correlation coefficient (R) is shown with the associated 95% CI. For testing the null hypothesis R=0, p<0.001. **C:** Recurrence probabilities estimated by multivariate logistic regression analyses (solid black line, dashed black line: 95% CI) from sequential PB measurements during dose de-escalation using the calculated intercept (left) and slope (right) as independent explanatory variables. Dots are individual intercept (left) and slope (right) values of patients who showed TFR (blue) or recurrence (red). The intercept is given in terms of molecular response and the slope in terms of increase in molecular response per month. Estimated ORs and associated 95% CIs describe the increase in the chance of losing TFR if intercept is increasing by one unit and if slope is increasing by 0.01 unit. BM: bone marrow, PB: peripheral blood, ND: not detectable, TFR: treatment-free remission, MNC: mononuclear cell, IQR: interquartile range, CI: confidence interval, OR: odds ratio, MMR: major molecular response, DMR: deep molecular response.

Univariate logistic regression shows that BCR::ABL1^IS^ levels, from either BM or PB taken at TKI de-escalation, were similarly predictive of TFR success at 36 months post-de-escalation (Suppl. Figures 4A, B). Thus, a benefit of BM over PB evaluations at the point of de-escalation was not shown.

Next, we used sequential PB BCR::ABL1^IS^ levels taken during TKI de-escalation to fit a linear regression model. The estimated regression parameters, intercept and slope, were tested as predictors of TFR success. In the univariate model, the intercept was associated with a higher odds ratio (OR) compared to a single BCR::ABL1^IS^ value from BM or PB at trial entry, as previously determined (Suppl. Figure 4C). In contrast, the regression slope did not show an increased OR, compared to the single BM or PB values at de-escalation (Suppl. Figure 4D). However, when using both the regression slope and intercept as independent predictors in a multivariate logistic regression model, the predictive value of the intercept was even more pronounced if adjusted for the slope effect, which itself remained predictive of TFR (Figure 1C, Suppl. Table 3).

Based on these findings, we used the slope and intercept to develop a risk prediction model for molecular recurrence. Using a comparative ROC analysis (see Suppl. Information), we classified DESTINY patients into three risk groups: *low, high* and *unclear* (Figure 2, Suppl. Figure 5A, B). For patients who could be classified into the *high* or *low risk* group, this model had high positive (PPV) and negative predictive values (NPV) of 100% and 85.2%, respectively, and a misclassification rate of only 10.8%. The difference in MRFS between the risk groups is illustrated in Figure 2B using stratified Kaplan–Meier curves.

**Figure 2.**
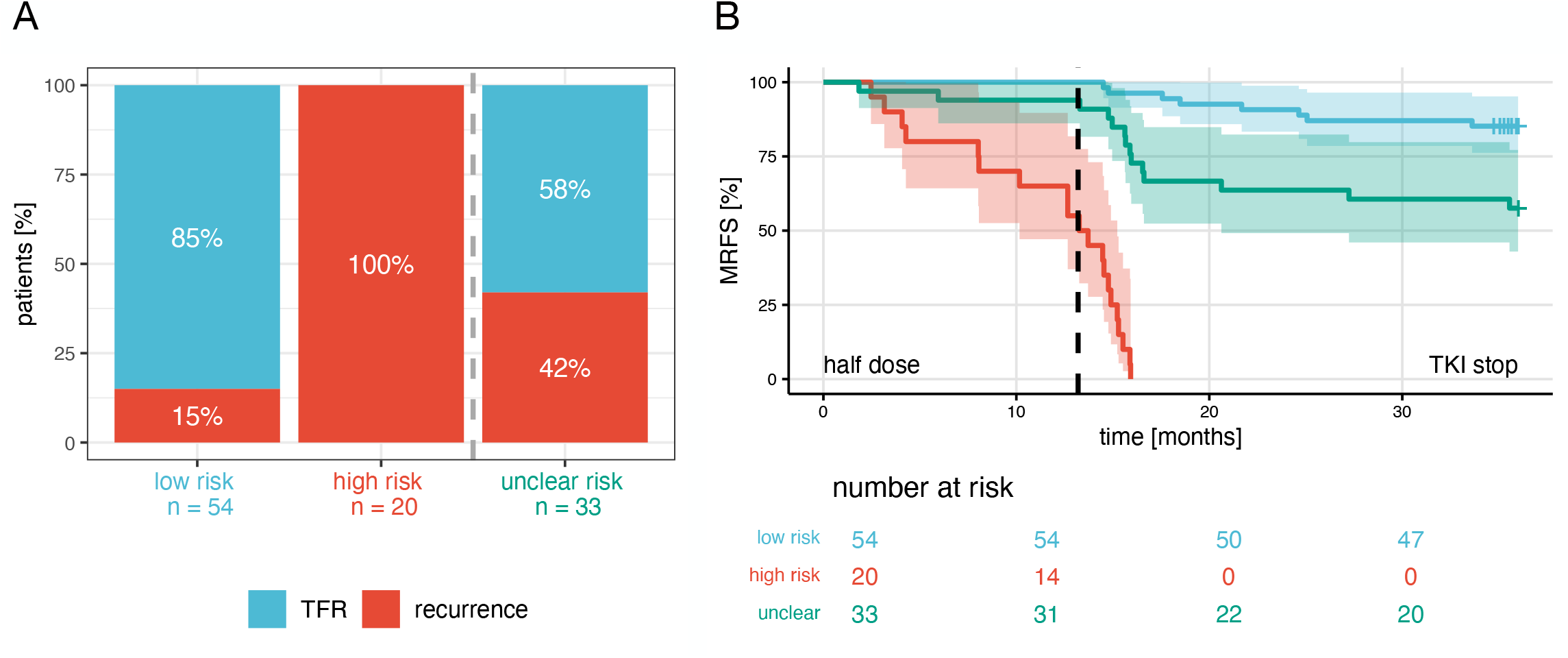
Risk of relapse following TKI de-escalation can be classified based on intercept and slope, estimated from sequential peripheral blood-based BCR::ABL1^IS^ measurements during TKI dose de-escalation. **A:** Proportion of patients in TFR or with recurrence events until the end of the 24-month follow-up period after TKI cessation in the three risk groups based on joined classification using intercept and slope. Absolute patient numbers given below the bars. **B:** Kaplan–Meier curve for MRFS (shaded areas: 95% confidence intervals), stratified according to the three risk classification groups; low, high and unclear. The dashed line represents the end of the de-escalation phase (12 months). The trial endpoint was set to 36 months, i.e., 24-month follow-up post-TKI cessation. Thus, patients were censored at last seen date before or at the end of trial. TFR: Treatment-free remission, MRFS: molecular recurrence-free survival, TKI: tyrosine kinase inhibitor.

In comparison to the slope and intercept evaluation, risk classification using single-timepoint BCR::ABL1 ^IS^ measurements from either BM or PB immediately prior to de-escalation were not as predictive of molecular recurrence, neither regarding their PPV nor NPV. Also, both models had higher rates of group misclassification (Suppl. Figure 6). Furthermore, adding these single timepoint BM or PB measurements to the multivariate (intercept + slope) model as independent predictors did not result in a significantly better model fit (Suppl. Table 4).

## Discussion

Our results demonstrate that TFR success can be more strongly predicted using the slope and intercept estimated by a linear regression from sequential BCR::ABL1^IS^ evaluation from PB during TKI de-escalation compared to a single BM or PB derived value determined prior to de-escalation. For 8 out of 10 patients who experienced recurrence during TKI de-escalation in DESTINY, a *high risk* classification was evident before the recurrence event; the remaining two patients were classified as *unclear* (Suppl. Fig. 7). These results indicate that a reliable risk classification is achievable after three months of TKI de-escalation, if ≥3 PB BCR::ABL1^IS^ measurements are available. If so, the BCR::ABL1^IS^ at the time of TKI de-escalation can more reliably be estimated by the regression intercept than by a single evaluation of either PB or BM samples.

In our analysis, there were very few undetectable BCR::ABL1^IS^ values from PB during TKI de-escalation, so these could be omitted without affecting the results. Thus, estimation of the regression slope and intercept is achievable for most patients with ≥3 PB BCR::ABL1^IS^ measurements, and more sophisticated measures would be necessary to estimate these predictive parameters for those patients with undetectable BCR::ABL1^IS^. The undetectable BCR::ABL1^IS^ values raise another important issue regarding the comparison of PB versus BM.

In 42 patients, BCR::ABL1^IS^ was detectable in PB, but not BM. A possible reason for this is that patients in DMR may only have scattered small pockets, if any, of *BCR::ABL1* positive cells within the BM, as suggested by imaging results for murine progenitor cells [10]. Therefore, obtaining a positive *BCR::ABL1* result is dependent on the area of the BM being sampled, whereas PB is assumed to provide a more homogenous *BCR::ABL1* signal. This observation supports the use of sequential PB-based BCR::ABL1^IS^ measurements as a more robust method to predict TFR success.

Within DESTINY, late molecular recurrences occurred beyond the trial endpoint of 24 months post-TKI cessation for n=5 patients (Suppl. Figure 8). In the presented analyses, these patients’ outcomes are classified as TFR, as recorded at the endpoint. Of these 5 patients, 3 were classified as *low risk*, and their BM and PB BCR::ABL1^IS^ values at study entry were relatively low and comparable to those of the other TFR patients. If these patients were analyzed as recurrences, the classification cut-off value for the regression slope would change considerably: while the misclassification rate among *high*/*low risk* patients would decrease from 10.8% to 6.7%, the number of patients with *unclear risk* would increase from 30.8% to 44%. This illustrates that the classification is dependent on the timing of recurrence. Accordingly, if the BCR::ABL1^IS^ slope remains positive, molecular recurrence will eventually occur; however, the timing of the recurrence depends quantitatively on the slope as well as on the intercept, which is effectively a robust estimate of the molecular response at the time of TKI de-escalation.

In conclusion, our results provide a strong rationale for a prospective clinical trial to test the suggested three-group risk classification, to improve TFR rates for *low risk* patients while minimizing the risk of molecular recurrence for *high risk* patients. During TKI de-escalation, the slope and intercept-based risk classification would be used to determine if or when a patient could discontinue TKI successfully, based on ≥ 3 sequential PB-based BCR::ABL1^IS^ measurements. Finally, these results show that there is no benefit from quantifying BCR::ABL1^IS^ from BM over PB for the prediction of TFR success, which is an important conclusion for patients.

## Acknowledgements

We would like to thank all patients and staff who supported the DESTINY clinical trial. The authors would also like to express their sincere gratitude to the laboratory technicians for their invaluable technical support throughout this research, and to Dr. Letizia Foroni for leading the Specialist Integrated Haematology Malignancies Diagnostic Service (SIHMDS), without whom this work would not have been possible.

## Funding

This study was funded by a Cancer Research UK Biomarker Award (CRCPJT/100006) and Blood Cancer UK Award (formerly Bloodwise; 11017). The DESTINY clinical trial was funded by Blood Cancer UK (13020). The study was supported by the Glasgow Adult Experimental Cancer Medicine Centre funded by Cancer Research UK.

## Competing Interests Statement

M.C. has received research funding from Cyclacel and Incyte, is/has been an advisory board member for Novartis, Incyte, Ascentage, Jazz Pharmaceuticals, Pfizer and Servier, and has received honoraria from Astellas, Novartis, Incyte, Pfizer, Janssen and Jazz Pharmaceuticals.

J.F.A has received research funding from Incyte and Pfizer, is/has been an advisory board member for Ascentage, Incyte, Novartis and Terns, and has received honoraria from Incyte and Novartis.

The remaining authors have no disclosures.

## Author Contributions

The study was conceived by MC, IR, HGJ, HWW, and REC. SDP and CH performed experiments. IR and AG designed the mathematical analysis strategy and AG performed the analysis. RY and IG provided support regarding data analysis/modelling. AH provided biobank support. SDP, AG, IR, IG and MC wrote the manuscript. JFA oversaw the centralized PB PCR testing. All authors commented on and approved the final manuscript.

## Ethical Approval

This study was conducted under the DESTINY clinical trial ethics approval from the NRES Committee North West and Liverpool East (13/NW/0265) and the Haematological Cell Research Bank Research (Tissue Bank Approval) from the West of Scotland REC 4 (20/WS/0066).

## Data Availability Statement

All data generated or analysed using PB-derived samples during this study are available as published in [9]. Researchers wishing access to the BM qRT-PCR data should contact the corresponding author.

## Notes

### Clinical Trial

NCT01804985

