## Supplemental Information for "BCR::ABL1 evaluation in bone marrow is no more predictive of TFR success than from peripheral blood"

**Supplemental Information Contents**

*(.pdf file)*

- **Clinical trial data:** Details of the DESTINY study and patient cohort.
- **Methods to measure BCR::ABL1$^{IS}$:** Technical description of methods of BCR::ABL1$^{IS}$ quantification.
- **Statistical methods:** Description of applied statistical analyses procedures including results (e.g., model parameters).
- **Supplemental Figures [1–8] and Tables [1–6].**
- **Supplemental references.**

**Clinical trial data**

All presented analyses are based on data from the DESTINY trial (NCT01804985; [S1, S2]), with n=174 chronic phase CML patients in stable remission. Of these, n=125 were in deep molecular remission (DMR) and n=49 in major molecular remission (MMR); n=148 were treated with imatinib, n=16 with nilotinib, and n=10 with dasatinib.

Clinical BCR::ABL1$^{IS}$ transcript data from the peripheral blood (PB) of DESTINY trial participants (n=174) were available at the trial entry immediately prior to de-escalation (as published in [S3]) and sequentially following this period. Additionally, mononuclear cells (MNCs) were isolated from bone marrow (BM) and PB immediately prior to TKI de-escalation. N=107 patients could be included in the final analyses (see Suppl. Fig. 1, Suppl. Table 2).

According to the DESTINY study protocol [S1,S2], molecular recurrence was defined as the first of two consecutive measurements reporting BCR-ABL1$^{IS}$ >0.1%. Furthermore, we considered n=3 additional recurrences reported in *case report forms,* in which the BCR-ABL1$^{IS}$ >0.1% threshold was not reached. This corresponds to final trial results posted on EUDRACT.

### Methods to measure BCR::ABL1$^{IS}$

Sample Processing and Cell Lysate Preparation

PB white blood cell lysates were prepared by selective lysis of red blood cells using the Puregene RBC Lysis Solution (Qiagen), followed by washing with phosphate-buffered saline (PBS) (Thermo Fisher Scientific). BM samples were processed to isolate mononuclear cells, which were subsequently preserved in RNAlater (Thermo Fisher Scientific) at –20°C. Prior to lysis, RNAlater was removed, and a lysis buffer consisting of Buffer RLT Plus (Qiagen), 2-mercaptoethanol (Sigma-Aldrich), and Reagent DX (Qiagen) was added to both PB and BM cell pellets. Homogenization was carried out using the TissueLyser II (Qiagen), and the resulting cell lysates were stored at –20°C until further processing.

RNA Extraction and cDNA Synthesis

Total RNA was extracted from PB and BM lysates using the QIAsymphony platform in conjunction with the QIAsymphony RNA Kit (Qiagen). Complementary DNA (cDNA) was synthesised from total RNA using the LunaScript™ RT SuperMix Kit (New England Biolabs), following the standard reverse transcription protocol.

Real-Time Quantitative PCR (RT-qPCR)

RT-qPCR was performed on the QuantStudio™ 5 Real-Time PCR System (Thermo Fisher Scientific) using a duplex assay targeting the *BCR::ABL1* fusion transcript (e13a2/e14a2) and the *ABL1* reference gene. Custom TaqMan™ primers and probes (Thermo Fisher Scientific; see Suppl. Table 1) were used in conjunction with the TaqMan™ Fast Advanced Master Mix (Thermo Fisher Scientific). Quantification was calibrated using the ERMAD623 *BCR-ABL* plasmid DNA (ERM®).

PCR reactions (20 µL total volume) contained 3 µL of cDNA and were run in triplicate under the following cycling conditions: polymerase activation at 95°C for 20 seconds, followed by 45 cycles of denaturation at 95°C for 3 seconds and annealing/extension at 60°C for 45 seconds (fast-mode). Results were expressed as *BCR::ABL1*/*ABL1* ratios and normalised to the international scale (IS) using a validated laboratory-specific conversion factor. A minimum of 10,000 *ABL1* copies was required for reliable *BCR::ABL1* quantification. Data analysis was performed using QuantStudio™ Design and Analysis Software v1.5.1 (Thermo Fisher Scientific).

### Statistical methods

*Molecular response evaluation*

To describe and model the BCR::ABL1$^{IS}$, we applied the *BCR::ABL1* to *ABL1* ratio converted to the international scale (*IS*), i.e., (*BCR::ABL1 / ABL1) · 100% · CF* = BCR::ABL1$^{IS}$, with the conversion factor of the particular lab (*CF*). Molecular response (MR) levels are given in terms of log reductions: $\log_{10}$[(*BCR::ABL1 / ABL1) · CF*] = $\log_{10}$[*BCR::ABL1*$^{IS}$]. BCR::ABL1$^{IS}$ below MR6 (limit of detection, LOD), i.e. BCR::ABL1$^{IS}$ $\leq$ 0.0001%, are denoted as not detectable. Undetectable measurements are only considered for the statistical analysis of BM and PB MNCs prior to TKI dose de-escalation. After TKI dose de-escalation, BCR::ABL1$^{IS}$ below the detection limit in PB are not considered for the statistical analysis [S3]. Please note that all analyses that include such undetectable values are highly dependent on the chosen LOD cut-off, particularly if it exceeds the smallest measurable value. In our analysis, MR6 is below the smallest measurable value and receives the lowest rank of all measurements. Therefore, all rank-based analyses are not dependent on this.

*Statistical analyses methods to analyse molecular response levels*

Using Mann–Whitney U-tests we compared the median molecular response between TFR and recurrence patients for BM and PB measurements, respectively (Fig. 1 A). To statistically test the trends of molecular responses (i.e. null hypothesis: equality of more than two groups) we applied Kruskal–Wallis tests (Suppl. Fig. 3).

For correlation analysis of molecular response levels, non-detectable values were excluded and the Spearman's correlation coefficient (R) with the associated 95% CI was estimated. The p-value is given for testing the null hypothesis: R=0.

*Linear regression for parameter estimation during TKI dose de-escalation (see also* [S3])

Time courses of the $\log_{10}$[BCR::ABL1$^{IS}$] values (denoted as $LRATIO(t)$) within the 12 months dose reduction period are described by a linear function depending on time $t$ (in months), i.e.:

$LRATIO(t) = b + \beta t$ with intercept parameter $b$ and slope parameter $\beta$, assuming normally distributed errors with constant variance $\sigma^2$. For each individual patient $i$, we applied a standard linear regression model (R-package: *stats*, function: *lm*()) to fit the $LRATIO(t)$ function, thereby obtaining estimates of the intercept $\hat{b}_i$ and the slope $\hat{\beta}_i$ along with an estimated residual variance $\hat{\sigma}_i^2$ on the log-scale. The fitting routine is only applied if at least three eligible observations are obtained during the dose reduction period. An observation is considered eligible when: 1) BCR::ABL1$^{IS}$ is detected (i.e. measurements are above the individual detection limit of the qRT-PCR) and 2) it occurred before a confirmed molecular recurrence (cited from [S3]). Finally, we included n=107 patients in the analysis which fulfil those criteria.

*<u>Logistic Regression</u>*

A logistic regression model was applied to analyze whether, besides the change of the BCR::ABL1$^{IS}$ values during the 12 months TKI dose de-escalation period (slope $\beta$), the estimated intercept *b* or the $\log_{10}$[BCR::ABL1$^{IS}$] level prior TKI dose de-escalation period $c$ (from BM) and $d$ (from PB) are predictive for the recurrence status. Technically, we transformed the individual slope parameter $\beta' = \beta \cdot 100$ to achieve better numerical robustness. In the **full models** we analyzed at maximum three predictors for the probability $\pi$ of the occurrence of molecular recurrence (R-package: *stats*, function: *glm*()). I.e., given the probability $\mathrm{P}(\text{recurrence} \mid \beta', b, c) = \pi(\beta', b, c)$, the logistic regression model is

$$\ln\left(\frac{\pi(\beta', b, c)}{1 - \pi(\beta', b, c)}\right) = \gamma_0 + \gamma_1\beta' + \gamma_2 b + \gamma_3 c$$

and given the probability $\mathrm{P}(\text{recurrence} \mid \beta', b, d) = \pi(\beta', b, d)$, it is

$$\ln\left(\frac{\pi(\beta', b, d)}{1 - \pi(\beta', b, d)}\right) = \gamma_0 + \gamma_1\beta' + \gamma_2 b + \gamma_4 d$$

with the offset $\gamma_0$ and the additive slopes $\gamma_i, i = \{1, 2, 3, 4\}$.

We applied Wald tests and profile likelihood confidence intervals to assess the statistical significance of the slope-effects.

*Model comparison and parameter selection*

All model fits and model characteristics are summarized in Suppl. Tab. 3 and 4.

In the first step, we applied univariate logistic regression with only one of the four variables ($\beta', b, c, d$) as predictors and compared these four models with the Akaike information criterion (AIC), looking for the most suitable model (i.e. with lowest AIC). The AIC is given by $\mathrm{AIC} = 2\mathrm{k} - 2\log(\mathrm{L})$, with $\mathrm{k}$ as the number of estimated parameters and $\mathrm{L}$ as the maximum likelihood of the particular model. The model with the slope $\beta'$ showed the lowest AIC (similar to [S3]). In the second step, we asked whether the individually estimated intercept improved the univariate model, and compared the univariate model of predictor slope $\beta'$ with the bivariate model of the predictor slope $\beta'$ and intercept $b$. Using the likelihood ratio test (LRT), we compared the two nested models (R-package: *stats*, function: *anova*()). The test statistic is given by $\mathrm{D} = -2 \cdot (\log \mathrm{L}_0 - \log \mathrm{L}_1)$, with $\mathrm{L}_0$ = likelihood of the smaller model and $\mathrm{L}_1$ = likelihood of the more complex model. $\mathrm{D} \sim \chi 2(\mathrm{df})$, where $\mathrm{df}$ is the number of additional parameters in the more complex model. The individually estimated intercepts $b$ improved the univariate model and we used the bivariate model as the reference model for the following steps.

In the third step, we asked whether molecular response levels prior TKI dose de-escalation period $c$ from BM or $d$ from PB improve the bivariate model. Therefore, we again used LRT to compare the nested models and could show that there is no further improvement by molecular response levels prior TKI dose de-escalation from BM or PB. Thus, the most predictive model is the bivariate model

$$\ln\left(\frac{\pi(\beta', b)}{1 - \pi(\beta', b)}\right) = \gamma_0 + \gamma_1 \beta' + \gamma_2 b.$$

*Identification of different response groups using ROC classification*

Due to the finding that the variables slope $\beta$ during dose de-escalation and intercept $b$, as well as molecular response level prior to TKI dose de-escalation from BM $c$ or PB $d$ are predictive as univariate predictors for the occurrence of molecular recurrence, we aimed to categorize the patients into clinically relevant groups. In order to identify a cut-off value that adequately separates the patients into risk groups, we used the approach of the receiver operating characteristic (ROC). The four predictors ($\beta', b, c, d$) were studied with respect to their ability to predict the molecular recurrence by analysing the cut-off-specific sensitivities and specificities in a ROC curve (Suppl. Fig. 5). Herein the sensitivity and specificity are given as $sens = tp/(tp + fn)$ and $spec = tn/(tn + fp)$, respectively, with $tp$ = true positive, $tn$ = true negative, $fp$ = false positive and $fn$ = false negative. From this, we can derive the maximum Youden-Index [S4], which maximizes the sum of sensitivity and specificity (Youden-Index: $J = sens + spec - 1$) as one possible cut-off value.

To assess the classification quality of the different parameters, we calculate the classification error as well as the positive (PPV) and negative (NPV) predictive value, the latter referring to the probability of recurrence in the *high risk* group and the probability of TFR in the *low risk* group, respectively (Suppl. Tab. 5). Mathematically, the classification error is defined as $cr = ((\mathrm{fp} + \mathrm{fn})/(tp + tn + fp + fn))$ and the 95% confidence interval $cr \pm 1.96 \cdot \sqrt{cr \cdot (1 - cr)/(tp + tn + fp + fn)}$, while the positive predictive value is given as $ppv = tp/tp + fp]$ with 95% confidence interval $ppv \pm 1.96 \cdot \sqrt{ppv\,(1 - ppv)/(tp + fp)}$ and the negative predictive value is given as $npv = tn/(tn + fn)]$ with 95% confidence interval $npv \pm 1.96 \cdot \sqrt{npv\,(1 - npv)/(tn + fn)}$.

Based on the classification, we estimated the odds ratios $OR = (tn \cdot tp)/(fn \cdot fp)$ and the 95% confidence interval $95\%CI = OR \cdot \exp\left(\pm 1.96 \cdot \sqrt{\frac{1}{tn} + \frac{1}{tp} + \frac{1}{fn} + \frac{1}{fp}}\right)$, indicating whether the risk of recurrence is increased for the *high risk* group compared to the low risk group (Suppl. Tab. 6).

According to the reference logistic model, which is most predictive for TFR success (i.e. with predictors slope and intercept), we further developed an extended risk classification using three risk groups: *low risk, high risk, unclear risk.* Patients with both low intercept and negative or low slope value (i.e. intercept < -2.8 and slope < 0.035) were classified as *low risk* (n=54, 50%), whereas patients with both high intercept and high slope value (i.e. high intercept > -2.8 and high slope > 0.035) were classified as *high risk* (n=20, 19%). The remaining patients with either high intercept or high slope value (i.e. intercept > -2.8 and slope < 0.035 or intercept < -2.8 and slope > 0.035) were separated into *unclear risk* (n=33, 31%) (Suppl. Tab. 5 and 6).

This 3-group-classification is just one option, aiming for trustworthy statements with respect to TFR prediction. Although providing very low misclassifications of high and low risk predictions, it comes at the cost of a cohort of patients with unclear risk. Note that the model predictions are highly dependent on how the cut-off values are chosen and which optimization criterion is used. This choice depends on the primary goal: minimizing false TFR prediction or minimizing false recurrence predictions. Whereas the described 3-group-classification balances sensitivity (avoiding failure to predict TFR-loss) and specificity (avoiding prediction of recurrence for "TFR patients"), it would also be possible to maximize either sensitivity or specificity alone, but at the expense of each another.

*Time to molecular recurrence*

In order to analyse the time to event data (in our case: from day 1 of de-escalation until occurrence of molecular recurrence), we estimate the molecular recurrence-free survival (MRFS) probability by a Kaplan-Meier estimator (R-package: *survival*). We stratified according to the three risk groups *low risk*, *high risk* and *unclear risk* based on the cut-off values for the individual slope and intercept parameter.

### Supplemental Figures

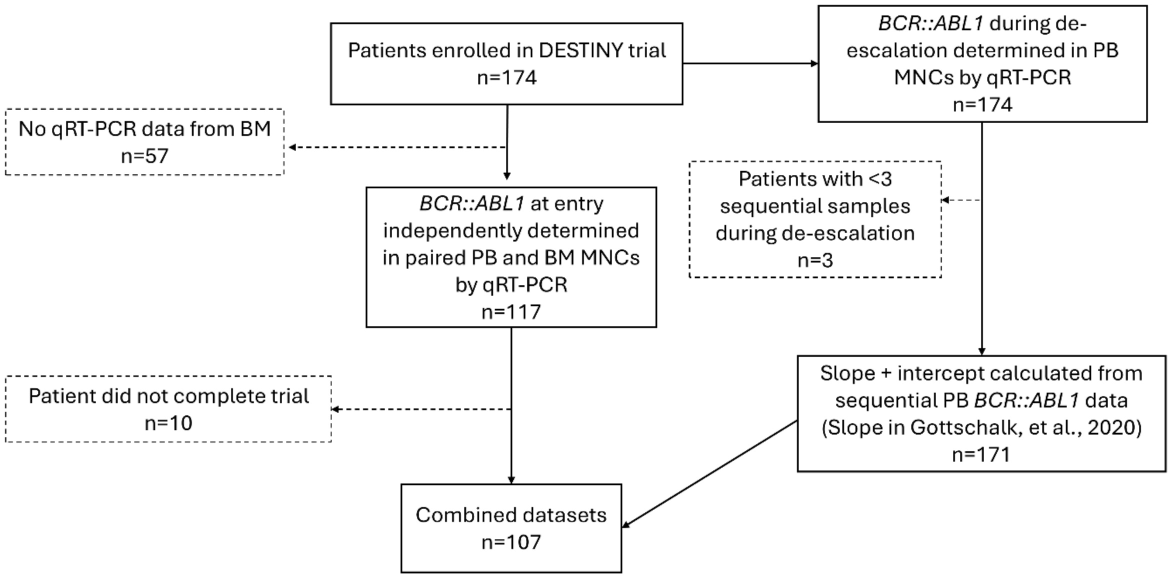


**Supplemental Figure 1. Sample selection process.** A subset of BM MNC samples from DESTINY trial participants were randomly selected for BCR::ABL1$^{IS}$ assessment. Samples were later excluded if qRT-PCR was unsuccessful or if there was no available data from paired PB MNC samples. Clinical BCR::ABL1$^{IS}$ transcript data from the PB of DESTINY trial participants (n=174) were available (as published in [S3]) at the trial entry point (immediately prior to de-escalation) and sequentially following this period; 3 patients had below the minimum requirement of 3 measurements for the regression and were excluded. The n=10 patients did not complete the study due to reasons other than molecular recurrence, including trial protocol violation, withdrawal of consent or unrelated mortality.

BM; bone marrow, PB; peripheral blood, MNC; mononuclear cell, TFR; treatment-free remission.

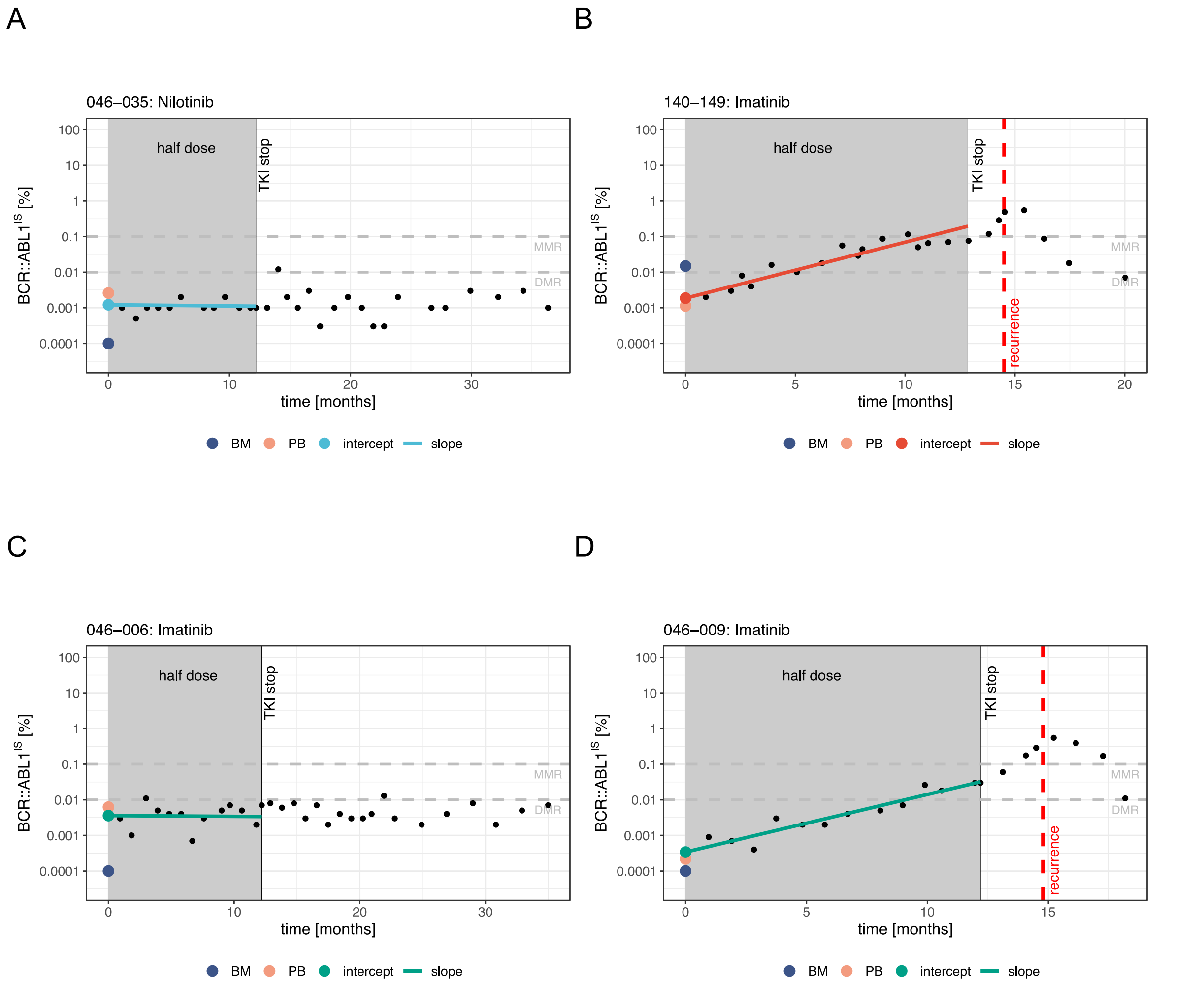


**Supplemental Figure 2: Examples of linear regression-based characterization of BCR::ABL1$^{IS}$ dynamics during TKI dose de-escalation according to risk groups.** Shown are representative examples of BCR::ABL1$^{IS}$ dynamics, classified as *low* (blue), *high* (red) or *unclear* (turquoise) risk, together with their slope and intercept (estimated by linear regression for *molecular response* levels in peripheral blood (PB) during dose de-escalation). Additionally, BCR::ABL1$^{IS}$, measured at a single time point immediately prior to TKI dose de-escalation in BM and PB is shown by dark blue and orange dots, respectively. **A:** Example patient with *low* risk classification, i.e. intercept < -2.8 and slope < 0.035 (blue dot and line). **B:** Example patient with *high* risk, i.e. high intercept > -2.8 and high slope > 0.035 (red solid line). **C**/**D:** Example patients with *unclear* risk (intercept + slope: turquoise dot and line): **C:** intercept > -2.8 and slope < 0.035; **D:** intercept < -2.8 and slope > 0.035. BCR::ABL1$^{IS}$ before TKI dose de-escalation are shown as deep blue (BM) and salmon (PB) dots; levels in PB after TKI dose de-escalation are shown as black dots. Dashed red line: time point of recurrence, if applicable.

BM: bone marrow, PB: peripheral blood, TKI: tyrosine kinase inhibitor, MMR: major molecular response, DMR: deep molecular response.

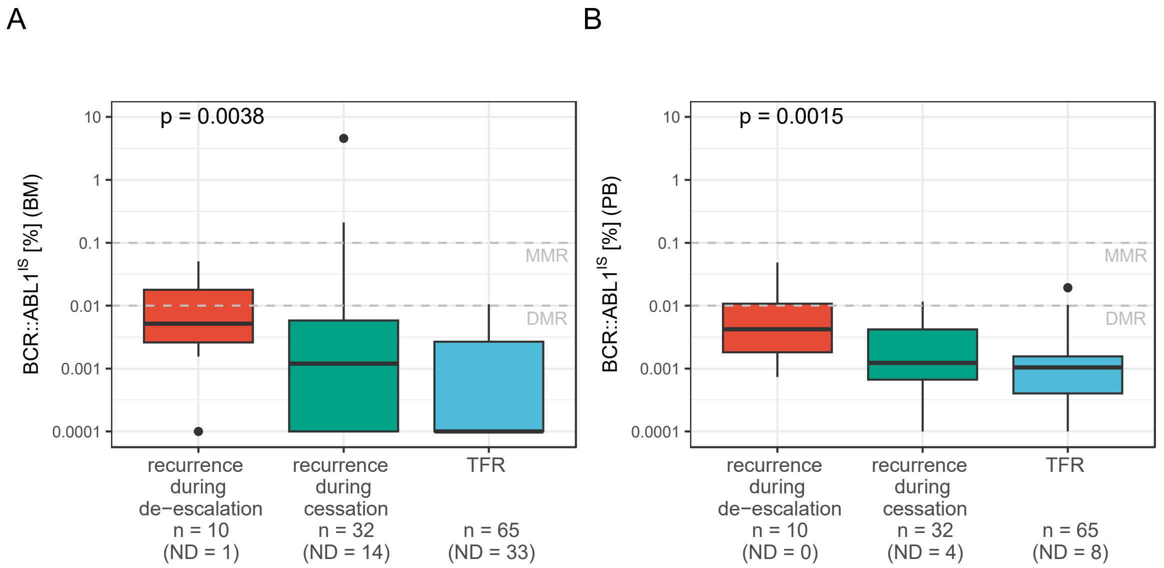


**Supplemental Figure 3: BCR::ABL1$^{IS}$ levels immediately prior to TKI dose de-escalation were highest in patients who subsequently relapsed during the de-escalation phase in both BM and PB.** Box plots, showing median (black line) and interquartile range (box), for BCR::ABL1$^{IS}$ as measured in BM **(A)** and PB **(B)** mononuclear cells, separated for the three outcome groups: recurrence during TKI de-escalation, i.e., study months 0-12 (red); recurrence during TKI cessation, i.e. study months 13–36 (turquoise); and TFR (no recurrence by 36 study months, blue). P-value refers to testing the statistical trends (i.e., decreasing BCR::ABL1$^{IS}$) using a Kruskal–Wallis test, i.e. testing for equality of all three groups.

BM: bone marrow, PB: peripheral blood, TKI: tyrosine kinase inhibitor, TFR: treatment-free remission, ND: not detectable, MMR: major molecular response, DMR: deep molecular response.

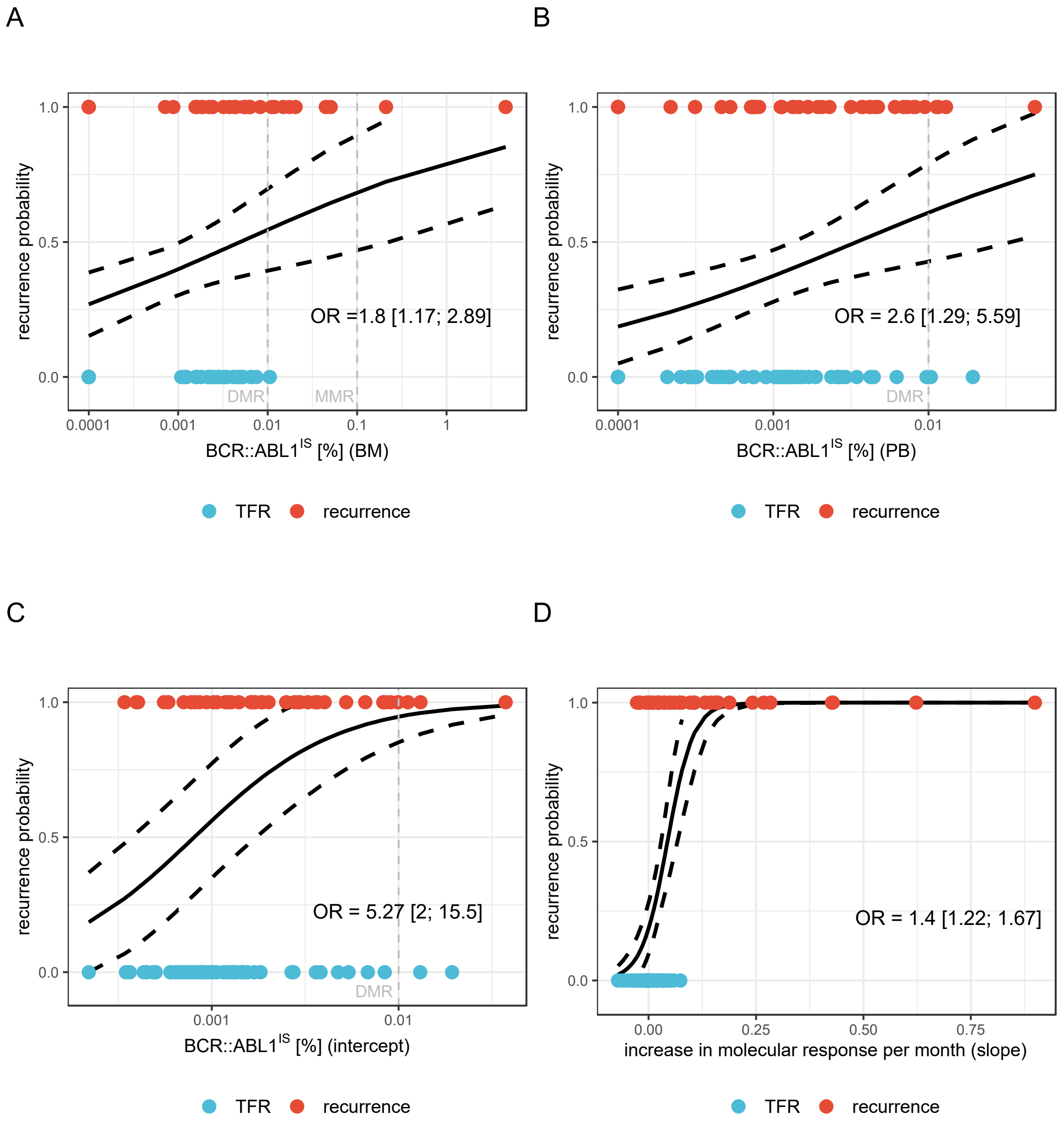


**Supplemental Figure 4: The PB-derived intercept value is more predictive of TFR outcome than single PB or BM BCR::ABL1$^{IS}$ measurements, based on univariate logistic regression.** Recurrence probabilities estimated by univariate logistic regression analyses (solid black line, dashed black lines: 95% CI) for the prediction of TFR. **A/B:** single time point immediately prior to TKI dose de-escalation; **A:** BM, **B:** PB**. C/D:** linear regression results, based on sequential PB levels during de-escalation. **C:** intercept, **D:** slope. Dots are referring to individual intercept (left) and slope (right) values of patients who showed TFR (blue) or recurrence (red). Estimated ORs and associated 95% CI describe the increase in the chance of losing TFR if BM, PB or intercept is increasing by one unit and if slope is increasing by 0.01 unit.

BM: bone marrow, PB: peripheral blood, TKI: tyrosine kinase inhibitor, TFR: treatment-free remission, OR: odds ratio, CI: confidence interval.

**A**

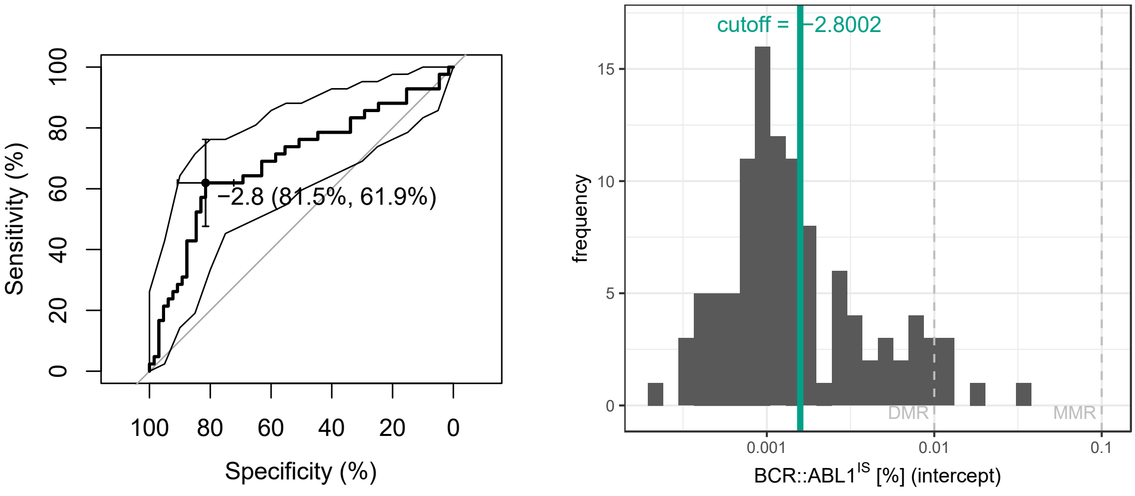


**B**

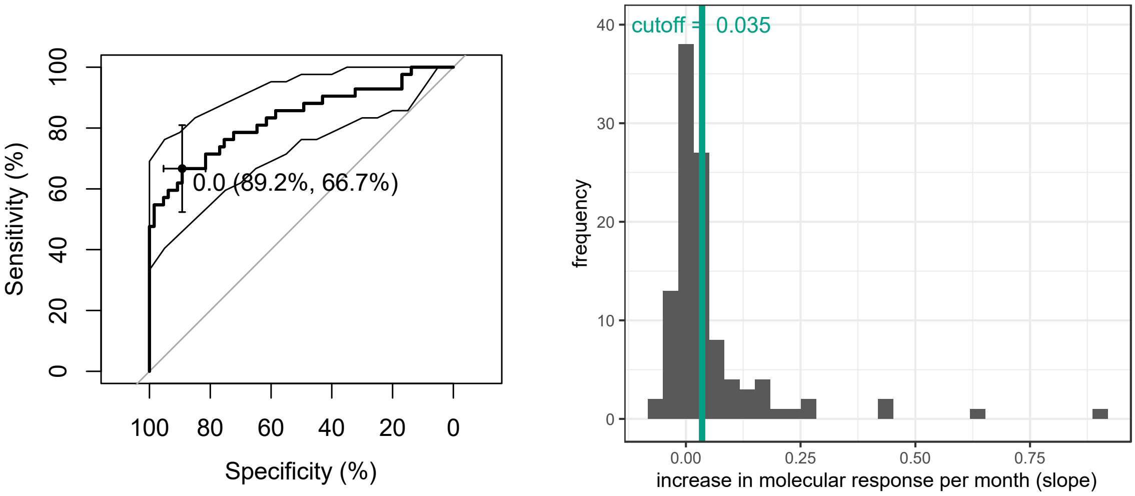


**Supplemental Figure 5: Classification cut-off determination by ROC analysis.** The intercept (**A**) and slope (**B**) values, determined by linear regression of sequential PB BCR::ABL1$^{IS}$ measurements during TKI dose de-escalation, were evaluated for their ability to predict TFR success using cut-off–specific sensitivities and specificities derived from receiver operating characteristic (ROC) analysis with associated 95% confidence intervals (turquoise band, left). Sensitivity and specificity are defined as: $sens = tp/(tp + fn)$ and $spec = tn/(tn + fp)$, with $tp$ = true positive, $tn$ = true negative, $fp$ = false positive and $fn$ = false negative. The optimal cut-off (highlighted in the ROC curve, left, and shown in the histogram, right) was determined using the maximum Youden Index: $J = sens + spec - 1$.

PB: peripheral blood, TKI: tyrosine kinase inhibitor, TFR: treatment-free remission, ROC: receiver operating characteristic.

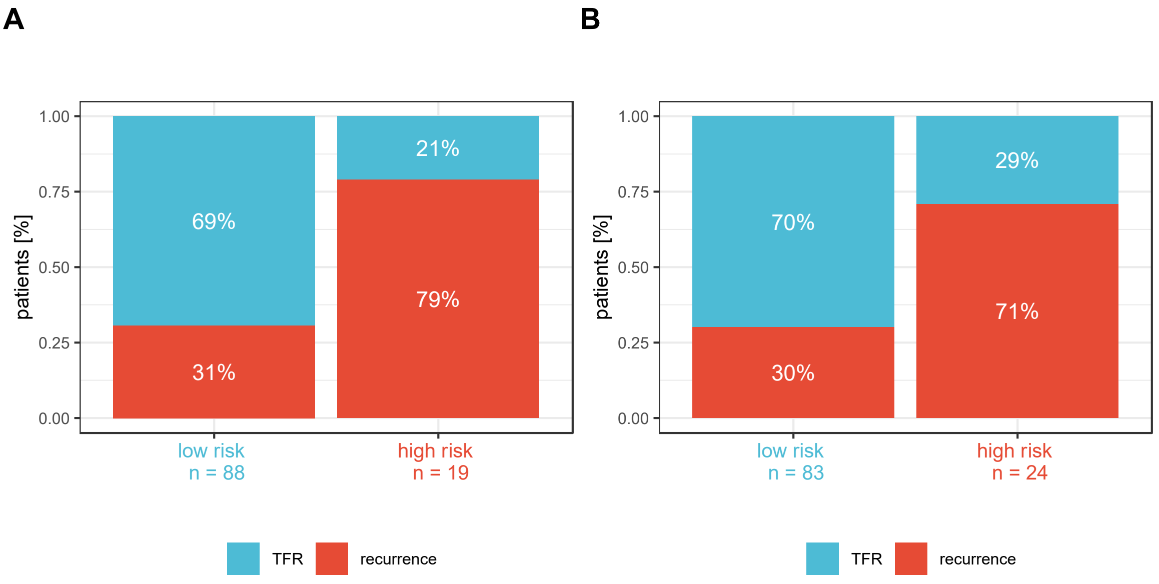


**Supplemental Figure 6. Risk classification based on single timepoint measurement before TKI de-escalation.** Outcomes (TFR vs. recurrence) at 24 months post TKI stop are shown for participants classified into *low* and *high* risk prediction groups, based on a previously determined cut-off (BM: -2.27, PB: -2.52) as measured in BM (**A**) or PB (**B**). Statistics associated with the predictive models are (**A**) BM: OR = 8.47 with 95% CI = [2.57; 27.92], PPV = 78,9% with 95% CI = [60.6%; 97.3%], NPV = 69.3% with 95% CI = [59.7%; 79.0%], misclassification: 29.0% with 95% CI [20.4; 37.6] and (**B**) PB: OR = 5.63 with 95% CI = [2.08; 15.27], PPV = 70,8% with 95% CI = [52.6%; 89.0%], NPV = 69.9% with 95% CI = [60.0%; 79.7%], misclassification: 29.9% with 95% CI [21.2: 38.6].

BM: bone marrow, PB: peripheral blood, TKI: tyrosine kinase inhibitor, TFR: treatment-free remission, OR: odds ratio, CI: confidence interval, PPV: positive predictive value, NPV: negative predictive value.

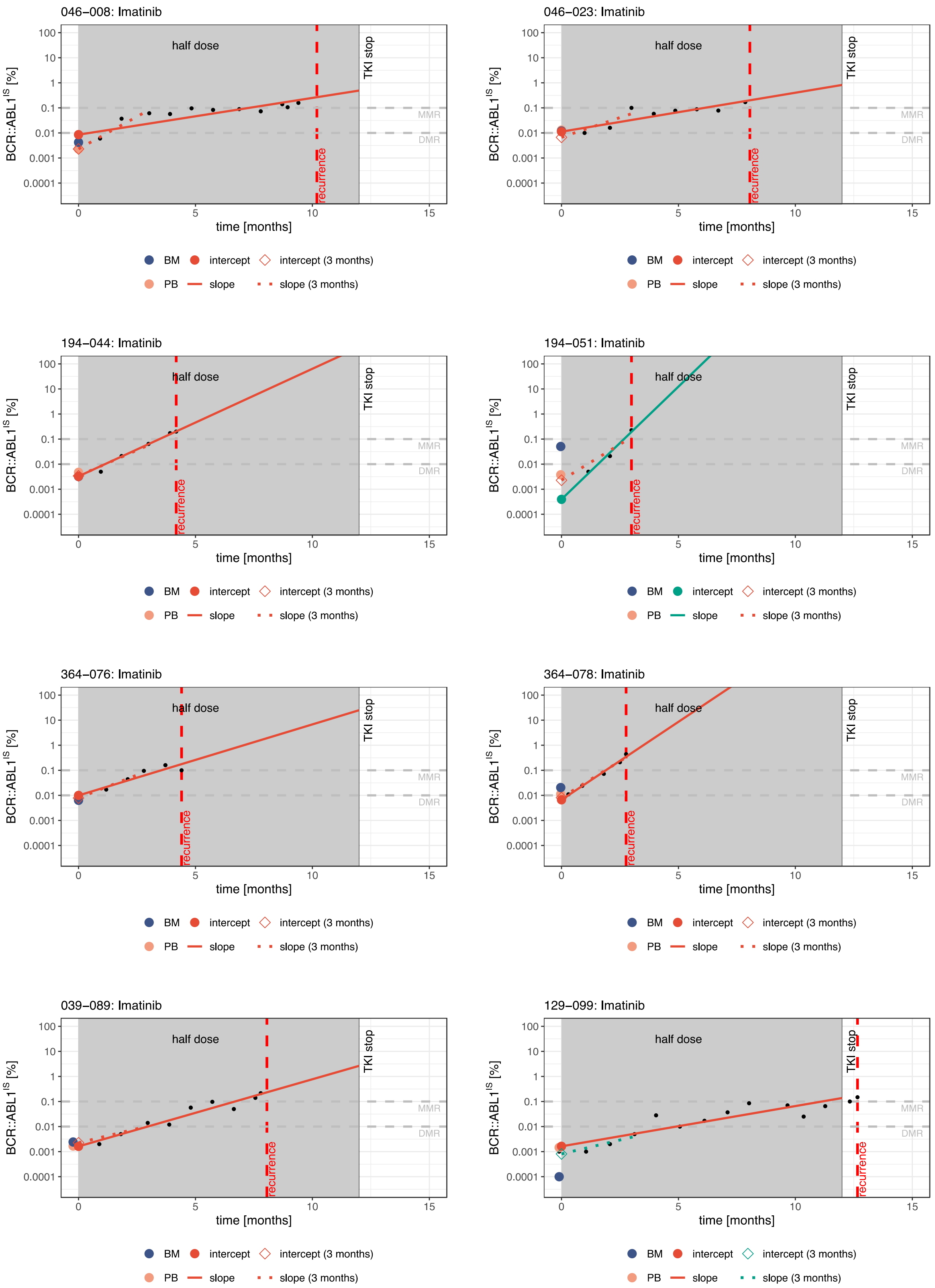
046−008: Imatinib
046−023: Imatinib
194−044: Imatinib
194−051: Imatinib
364−076: Imatinib
364−078: Imatinib
039−089: Imatinib
129−099: Imatinib
half dose
TKI stop
recurrence
MMR
DMR
BCR::ABL1$^{IS}$ [%]
100
10
1
0.1
0.01
0.001
0.0001
0
5
10
15
time [months]
BM
intercept
intercept (3 months)
PB
slope
slope (3 months)

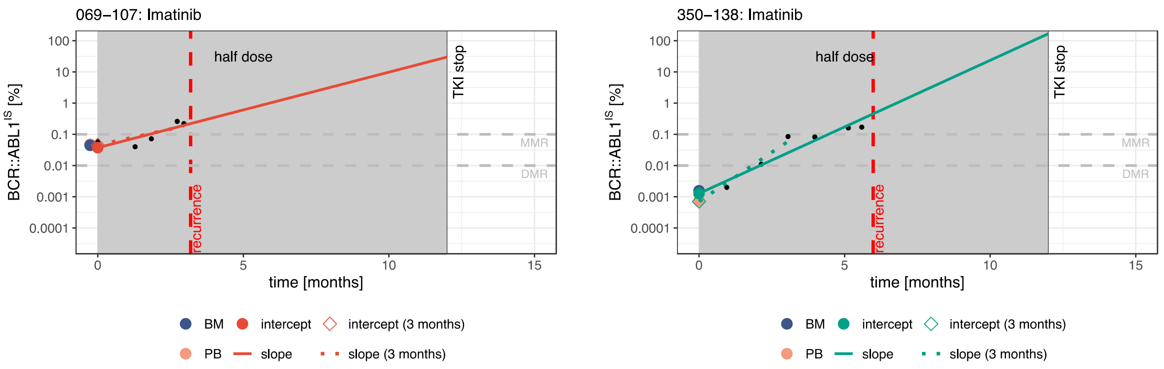


**Supplemental Figure 7. BCR::ABL1$^{IS}$ dynamics of all patients with recurrence within the tyrosine kinase inhibitor dose de-escalation phase.** Shown are the BCR::ABL1$^{IS}$ dynamics for patients with recurrence before TKI stopping (dynamics from all other patients can be obtained from the authors upon request), classified as *high* (i.e. high intercept > -2.8 and high slope > 0.035, red) or *unclear* (i.e. intercept < -2.8 and slope > 0.035, turquoise) risk, together with slope and intercept. BCR::ABL1$^{IS}$ levels before TKI dose de-escalation are shown as deep blue (BM) and salmon (PB) dots; BCR::ABL1$^{IS}$ levels in PB after TKI dose de-escalation are shown as black dots. Vertical dashed red line: Time point of recurrence. The slope and intercept estimates have been obtained from measurements of the first three months after TKI dose de-escalation, only (dotted increasing lines and same diamond) and from all measurements after TKI dose de-escalation (solid increasing lines and same colored dot). The uncertainty in risk classification (i.e., *unclear* risk as shown by turquoise slope and intercept) results from a rather low intercept (below MR5), such that even the large slope does not allow a *high* risk classification according to the rules described.

BM: bone marrow, PB: peripheral blood, TKI: tyrosine kinase inhibitor, MR5: molecular response level 5.

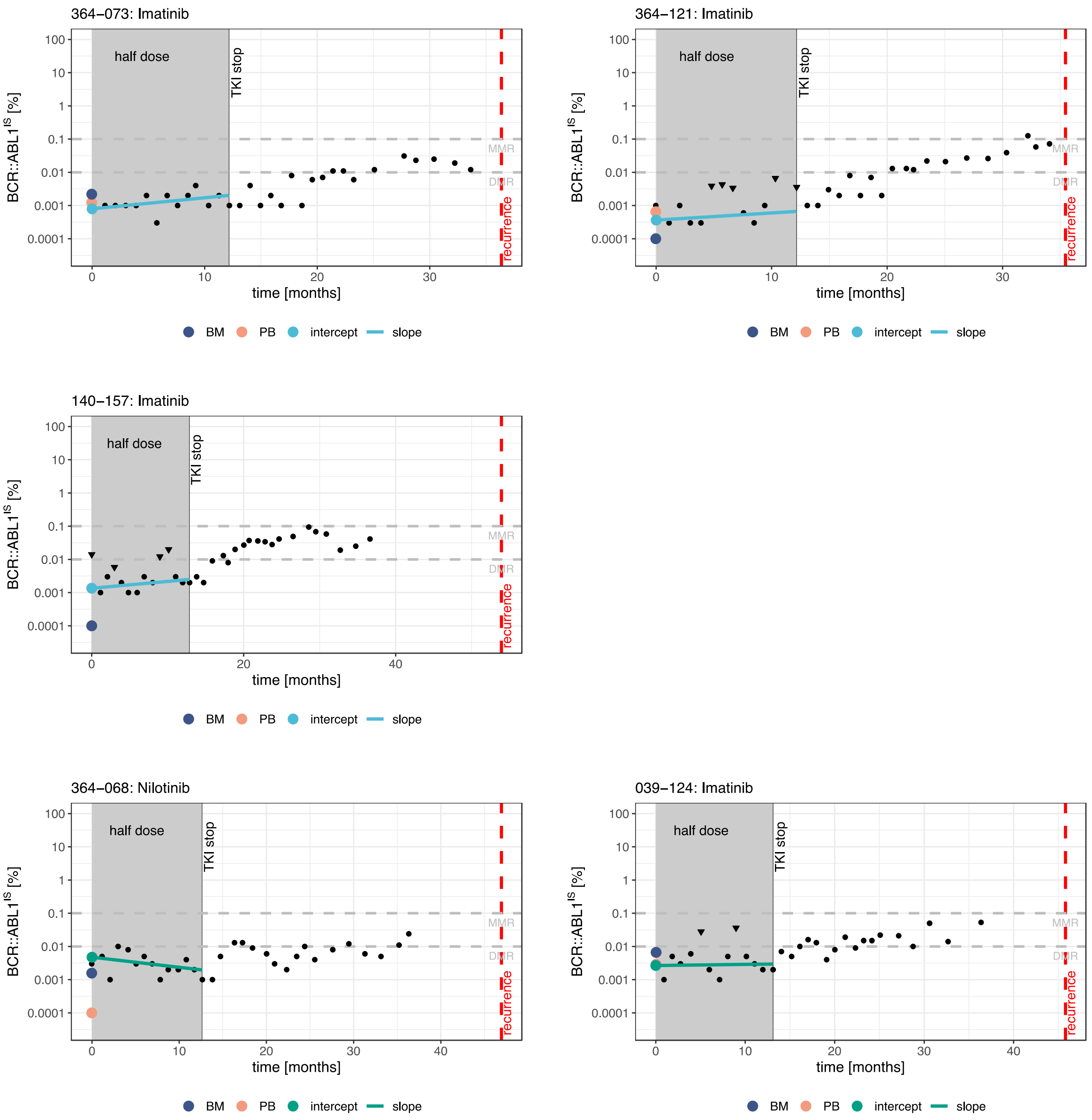


**Supplemental Figure 8. BCR::ABL1$^{IS}$ dynamics of all patients who experienced recurrence after the DESTINY trial endpoint, i.e. more than 24 months post TKI stop.** Explanation of symbols: see Suppl. Figure 7. Additionally, undetectable BCR::ABL1$^{IS}$ levels are shown as triangles. Measurements after 24 months post TKI stop are missing but the time points of recurrence are available (dashed red line). Risk classification calculated according to the assumption that these patients did stay in TFR, as they did within the 24-month post-TKI cessation follow-up period. These cases illustrate the dependence of the risk classification (including the cut-off choice) on the follow-up time that is considered to evaluate TFR / loss of TFR. However, BM and PB molecular response levels prior TKI dose de-escalation (light blue and purple dots) are low (BM: median [IQR] = -2.8 [-4; -2.6], PB: -3.2 [-4; -2.9]) and comparable to those of other TFR patients (BM: median [IQR] = -4 [-4; -2.59]; PB: -2.98 [-3.44; -2.81]).

BM: bone marrow, PB: peripheral blood, TKI: tyrosine kinase inhibitor, TFR: treatment-free remission, IQR: interquartile range.

### Supplemental Tables

| Assay | Primer/Probe Name | Gene [Exon] | Sequence (5'-3') | Final Concentrations |
|---|---|---|---|---|
| *BCR::ABL1* | ENF501 | *BCR [13]* | TCCGCTGACCATCAAYAAGGA | 300nM |
| | ENR561 | *ABL1 [2]* | CACTCAGACCCTGAGGCTCAA | 300nM |
| | FAM-ENP541-MGB | *ABL1 [2]* | FAM-CCCTTCAGCGGCCAGT-MGB | 100nM |
| *ABL1* | ENF1003 | *ABL1 [2]* | TGGAGATAACACTCTAAGCATAACTAAAGGT | 150nM |
| | ENR1063 | *ABL1 [3]* | GATGTAGTTGCTTGGGACCCA | 150nM |
| | VIC-ABL1043-MGB | *ABL1 [3]* | VIC-CATTTTTGGTTTGGGCTTC-MGB | 200nM |

**Supplemental Table 1. Primer and Probe Sequences and Final Concentrations.**

The primers, probes and final concentrations utilised in the duplex RT-qPCR reactions are those published [S5].

| Subset for PCR analyses (n=107) | | | |
|---|---|---|---|
| | | Recurrence | TFR |
| Sex | | | |
| | Male n= (%) | 20 (47.6) | 38 (58.5) |
| | Female n= (%) | 22 (52.4) | 27 (41.5) |
| Age at trial entry (years) | | | |
| | Median (range) | 57.5 (27-84) | 61 (28-81) |
| *BCR::ABL1* Group | | | |
| | MMR n= (%) | 25 (59.5) | 11 (16.9) |
| | DMR n= (%) | 17 (40.1) | 54 (83.1) |
| Transcript | | | |
| | e13a2 n= (%) | 11 (26.2) | 13 (20.0) |
| | e14a2 n= (%) | 21 (50.0) | 26 (40.0) |
| | Mixed n= (%) | 6 (14.3) | 13 (20.0) |
| | Unknown n= (%) | 4 (9.5) | 13 (20.0) |
| TKI at trial entry | | | |
| | IM n= (%) | 38 (90.5) | 54 (83.1) |
| | DAS n= (%) | 3 (7.1) | 3 (4.6) |
| | NIL n= (%) | 1 (2.4) | 8 (12.3) |

**Supplemental Table 2. Demographics of DESTINY trial participants.** Demographic data from the subset of trial participants from whom samples were used in this study (n=107). Percentages are within each outcome group.

DAS: dasatinib, DMR: deep molecular response, IM: imatinib, MMR: major molecular response, NIL: nilotinib, TFR: treatment-free remission, TKI: tyrosine kinase inhibitor.

| model | Coefficient | Estimate | Std.Error | p-value |
|---|---|---|---|---|
| A) BM | (offset) | 1.3601 | 0.7257 | |
| | BM | 0.5893 | 0.2305 | 0.0106 |
| | residual std. deviation σ | 136.19 | | |
| B) PB | (offset) | 2.3548 | 1.0902 | |
| | PB | 0.9561 | 0.3705 | 0.0099 |
| | residual std. deviation σ | 136.01 | | |
| C) intercept | (offset) | 4.2205 | 1.4643 | |
| | intercept | 1.6617 | 0.5178 | 0.0013 |
| | residual std. deviation σ | 131.48 | | |
| D) slope | (offset) | -1.46448 | 0.30903 | |
| | slope | 0.33360 | 0.07878 | <0.001 |
| | residual std. deviation σ | 94.16 | | |
| E) intercept + slope | (offset) | 5.9520 | 2.1859 | |
| | intercept | 2.6253 | 0.7830 | <0.001 |
| | slope | 0.4163 | 0.1023 | <0.001 |
| | residual std. deviation σ | 80.27 | | |
| F) intercept + slope + BM | (offset) | 5.97966 | 2.34426 | |
| | BM | 0.01107 | 0.33766 | 0.9738 |
| | intercept | 2.62252 | 0.78728 | <0.001 |
| | slope | 0.41608 | 0.10244 | <0.001 |
| | residual std. deviation σ | 80.27 | | |
| G) intercept + slope + PB | (offset) | 5.7524 | 2.2257 | |
| | PB | -0.5444 | 0.5929 | 0.3585 |
| | intercept | 3.1388 | 0.9853 | 0.0014 |
| | slope | 0.4391 | 0.1098 | <0.001 |
| | residual std. deviation σ | 79.44 | | |

**Supplemental Table 3. Model parameters from all logistic regression models applied.** Coefficient estimates and corresponding standard deviations of logistic regression models for TFR success together with p-value of the Wald significance test for predictor effects: **A)** Univariate model with single BCR::ABL1$^{IS}$ from bone marrow (BM) taken immediately prior to de-escalation as predictor, **B)** univariate model with single BCR::ABL1$^{IS}$ from peripheral blood (PB) taken immediately prior to de-escalation as predictor, **C)** univariate model with intercept (estimated by linear regression of multiple BCR::ABL1$^{IS}$ from PB during de-escalation) as a predictor, **D)** univariate model with slope (estimated by linear regression of multiple BCR::ABL1$^{IS}$ from PB during de-escalation) as predictor, **E)** bivariate model with slope and intercept (estimated by linear regression of multiple BCR::ABL1$^{IS}$ from PB during de-escalation) as predictors, **F)** combination of model A) and D) and **G)** combination of model B) and D). The parameters BM, PB, slope and intercept are reported in terms of BCR::ABL1$^{IS}$ level. The variable slope was multiplied by 100 for better interpretation and robust fitting.

BM: bone marrow, PB: peripheral blood, TFR: treatment-free remission.

| **model** | **AIC** | **p-value** (compared to reference model) |
|---|---|---|
| **Univariate models** | | |
| A) BM | 140.19 | |
| B) PB | 140.01 | |
| C) intercept | 135.48 | <0.001 |
| D) slope | 98.16 | <0.001 |
| **Multivariate models** | | |
| **E) intercept + slope (reference)** | **86.27** | |
| F) intercept + slope + BM | 88.27 | 0.9738 |
| G) intercept + slope + PB | 87.44 | 0.3633 |

**Supplemental Table 4. Comparison of logistic regression models.** AIC of all logistic regression models for TFR success of Suppl. Table 3. p-values refer to pairwise model comparisons with model E (intercept + slope) as reference model using likelihood ratio test. Comparison of all univariate models, does not show any significant differences. Model E (intercept + slope), which is significantly more appropriate than the univariate models C (intercept) and D (slope) shows lowest AIC among all fitted models. Extending model E by either BM or PB values measured immediately prior to TKI dose de-escalation does not results in a significant change of the model fit.

BM: bone marrow, PB: peripheral blood, TFR: treatment-free remission, TKI: tyrosine kinase inhibitor, AIC: Akaike information criterion.

| parameter | Cut-off value | number of *high risk* patients n (%) | classification error [95%CI] | ppv % [95%CI] | npv % [95%CI] |
|---|---|---|---|---|---|
| BM | -2.27 | 19 (17.8) | 29.0 [20.4; 37.6] | 78.9, [60.6; 97.3] | 69.3 [59.7; 79.0] |
| PB | -2.52 | 24 (22.4) | 29.9 [21.2; 38.6] | 70,8 [52.6; 89.0 | 69.9 [60.0; 79.7] |
| intercept | -2.8 | 38 (35.5) | 26.2 [17.8; 34.5] | 68.4 [53.6, 83,2] | 76.8 [66.9; 86.8] |
| slope | 0.035 | 35 (32.7) | 20.2 [12.6; 27.7] | 77.8% [64.2; 91.4] | 80.8 [71.8; 89.9] |
| slope + Intercept* | 0.035 -2.8 | 20 (27.0) | 10.8 [3.7; 17.9] | 100 | 85.2 [75.7; 94.7] |

**Supplemental Table 5: Characteristics of the cut-off values of the different parameters and the reference model (n=107).**

**low* and *high* risk groups only (n=74)

BM: bone marrow, PB: peripheral blood, PPV: positive predictive value, NPV: negative predictive value.

| | TFR | | | recurrence | | | |
|---|---|---|---|---|---|---|---|
| | Low risk | High risk | Unclear risk | Low risk | High risk | Unclear risk | OR [95%CI] |
| BM | 61 | 4 | | 27 | 15 | | 8.47 [2.57; 27.92] |
| PB | 58 | 7 | | 25 | 17 | | 5.63 [2.08; 15.27] |
| intercept | 53 | 12 | | 16 | 26 | | 7.18 [2.97; 17.36] |
| slope | 58 | 7 | | 14 | 28 | | 16.57 [6.02; 45.65] |
| slope + intercept | 46 | 0 | 19 | 8 | 20 | 14 | - |

**Supplemental Table 6: Contingency table showing the number of patients classified for recurrence status and the particular parameter(s) (n=107).**

BM: bone marrow, PB: peripheral blood, TFR; treatment-free remission, OR: odds ratio, CI: confidence interval.
